# Risk factors for severe outcomes of COVID-19: a rapid review

**DOI:** 10.1101/2020.08.27.20183434

**Authors:** Aireen Wingert, Jennifer Pillay, Michelle Gates, Samantha Guitard, Sholeh Rahman, Andrew Beck, Ben Vandermeer, Lisa Hartling

## Abstract

**Background:** Identification of high-risk groups is needed to inform COVID-19 vaccine prioritization strategies in Canada. A rapid review was conducted to determine the magnitude of association between potential risk factors and risk of severe outcomes of COVID-19.

**Methods:** Methods, inclusion criteria, and outcomes were prespecified in a protocol that is publicly available. Ovid MEDLINE(R) ALL, Epistemonikos COVID-19 in L·OVE Platform, and McMaster COVID-19 Evidence Alerts, and select websites were searched to 15 June 2020. Studies needed to be conducted in Organisation for Economic Co-operation and Development countries and have used multivariate analyses to adjust for potential confounders. After piloting, screening, data extraction, and quality appraisal were all performed by a single reviewer. Authors collaborated to synthesize the findings narratively and appraise the certainty of the evidence for each risk factor-outcome association.

**Results:** Of 3,740 unique records identified, 34 were included in the review. The studies included median 596 (range 44 to 418,794) participants with a mean age between 42 and 84 years. Half of the studies (17/34) were conducted in the United States and 19/34 (56%) were rated as good quality. There was low or moderate certainty evidence for a large (≥2-fold) association with increased risk of hospitalization in people having confirmed COVID-19, for the following risk factors: obesity class III, heart failure, diabetes, chronic kidney disease, dementia, age over 45 years (vs. younger), male gender, Black race/ethnicity (vs. non-Hispanic white), homelessness, and low income (vs. above average). Age over 60 and 70 years may be associated with large increases in the rate of mechanical ventilation and severe disease, respectively. For mortality, a large association with increased risk may exist for liver disease, Bangladeshi ethnicity (vs. British white), age over 45 years (vs. <45 years), age over 80 years (vs. 65-69 years), and male gender in those 20-64 years (but not older). Associations with hospitalization and mortality may be very large (≥5-fold increased risk) for those aged over 60 years.

**Conclusion:** Among other factors, increasing age (especially >60 years) appears to be the most important risk factor for severe outcomes among those with COVID-19. There is a need for high quality primary research (accounting for multiple confounders) to better understand the level of risk that might be associated with immigration or refugee status, religion or belief system, social capital, substance use disorders, pregnancy, Indigenous identity, living with a disability, and differing levels of risk among children.

**PROSPERO registration:** CRD42020198001

What is already known
- The novel nature of COVID-19 means that in many countries there are currently no pre-determined priority groups for the receipt of the eventual COVID-19 vaccine(s).
- Primary research is rapidly emerging, but consensus on who might be at increased risk of severe outcomes from COVID-19 has not been established.

What this study adds
- This rapid review shows that advancing age (≥45 years and especially ≥60 years) may be the most important risk factor for hospitalization and mortality from COVID-19. Other important risk factors for severe disease identified by this review include several pre-existing chronic conditions (class III obesity, heart failure, diabetes, chronic kidney disease, liver disease, dementia), male gender, Black race/ethnicity (vs. non-Hispanic white), Bangladeshi ethnicity (vs. British white), low income (vs. high), and homelessness.

## INTRODUCTION

Novel coronavirus disease 2019 (COVID-19) is an infectious respiratory disease caused by the newly identified Severe Acute Respiratory Syndrome-Coronavirus-2 (SARS-CoV-2),[1] which reached worldwide pandemic status in early March 2020. [2] As of August 24, there were over 23 million confirmed cases worldwide and 800,000 deaths attributed to the virus. [3] Most people who develop COVID-19 will experience mild-to-moderate illness primarily affecting the respiratory system and recover at home. [4] In more severe cases, patients may require specialized care (e.g., admission to hospital and/or intensive care unit [ICU], assisted ventilation)[5] as the disease can progress to respiratory failure and/or affect multiple organ systems.[4] Though new primary research is emerging rapidly, the evidence is fragmented and consensus on who might be at increased risk of severe outcomes from COVID-19 has not been established.

Given the rapid spread of COVID-19 since its first emergence in late 2019, and potential for severe illness (including death), the development of a preventive vaccine has become a global priority. [6] COVID-19 vaccine development has been progressing at an unprecedented pace. Once a successful COVID-19 vaccine candidate becomes available, the initial vaccine supply is not expected to be sufficient to cover the entire population right away. Therefore, there is an urgent need to plan for the efficient, effective, and equitable allocation of eventual COVID-19 vaccines when limited initial vaccine supply will necessitate recommendations for the vaccination of certain groups earlier than others. Due to the novel nature of COVID-19, these groups for early vaccination have not yet been established.[7]

The National Advisory Committee on Immunization (NACI) is an expert advisory body that provides advice on the use of vaccines in Canada. [8] At the time of writing, NACI is developing interim guidance on priority pandemic immunization strategies for COVID-19 vaccination when initial vaccine supply is limited.[7] To inform this guidance, NACI is using its recently published Ethics, Equity, Feasibility and Acceptability (EEFA) Framework[9] to ensure these factors are systematically and comprehensively considered. One of the evidence informed tools that make up this framework is the “Equity Matrix” which has adapted the PROGRESS-Plus model of health determinants and outcomes[10] to ensure important vaccine-specific equity factors are explicitly included. The resulting “P^2^ROGRESS And Other Factors” framework includes a range of biological and social factors that likely contribute to inequities in health outcomes across population groups, but it is not yet clear how each factor might apply to COVID-19 outcomes. A discussion on the use of this Equity Matrix, with evidence from this rapid review, as a critical tool to guide the ethically just allocation of scarce resources is published elsewhere. [11]

With the aim of providing timely, evidence-informed guidance on pandemic vaccine prioritization, NACI required a rigorous and expedited synthesis of the available evidence on population groups who are at increased risk of severe illness and mortality as a result of COVID-19. Responding to this need, we conducted a rapid review to determine the magnitude of association between “P^2^ROGRESS And Other Factors” and risk of severe outcomes of COVID-19.

## METHODS

### Review Approach

The urgent need for empiric evidence to inform the prioritization of pandemic immunization strategies in Canada necessitated a rapid but rigorous approach to synthesizing the currently available data. Therefore, we performed a rapid review informed by traditional systematic review methodology, [12] with several modifications to allow for the evidence to be synthesized on an expedited timeline (e.g., single reviewer for study selection, data extraction, and assessment of risk of bias) and focusing on studies having high applicability to Canada (e.g., countries with universal health care system)

NACI’s High Consequence Infectious Disease Vaccine Working Group was consulted to develop and refine the scope of the review (i.e., priority population(s), risk condition(s)/factor(s), and outcomes of interest), but was not involved in the conduct of the review. The working group was not involved in selection of studies nor the synthesis of findings.

The review was conducted following an *a-priori* protocol (PROSPERO #CRD42020198001). Because there is not yet formal guidance on the reporting of rapid reviews, reporting adheres to the Preferred Reporting Items for Systematic Reviews and Meta-Analyses (PRISMA).[13]

### Literature Search

A health sciences librarian searched Ovid MEDLINE(R) ALL on 15 June 2020 using concepts related to COVID-19, P^2^ROGRESS And Other Factors, and severe outcomes (Supplemental File). The search was limited to studies published in English or French in 2020. Additionally, the search was limited to populations in countries that are members of the Organisation for Economic Cooperation and Development (OECD),[14] in an effort to include studies of highest relevance to the Canadian context. Editorials and letters were excluded. We supplemented the Medline search by hand-searching Epistemonikos COVID-19 in L^OVE Platform (https://app.iloveevidence.com/topics) and McMaster COVID-19 Evidence Alerts (https://plus.mcmaster.ca/COVID-19/) for relevant prognosis or aetiology studies up to 12 June 2020. A hand-search of relevant websites recommended by the NACI working group was also undertaken, as well as continual surveillance for publication of eligible pre-prints located by the search. Searches were exported to an Endnote Library (X9, Clarivate Analytics, Philadelphia, PA) and duplicates removed.

### Eligibility Criteria

We included studies published in English or French since 1 January 2020 that reported on the magnitude of association between potential P^2^ROGRESS And Other Factors and severe outcomes of COVID-19 (Supplemental File). Eligible populations, in order of priority, were people (a) from a general/community sample, (b) with COVID-19 confirmed (by laboratory testing or epidemiologic linkage), (c) hospitalized with COVID-19, and d) with a risk factor of interest. To ensure relevance to the Canadian context, studies had to be conducted in OECD countries;[14] we included studies from countries that do not provide universal (or near universal) coverage for core medical services (i.e., Chile, Greece, Mexico, Poland, the Slovak Republic, and the United States)[15] but considered these to be less applicable to the Canadian context when interpreting the findings.

The exposures of interest were any P^2^ROGRESS And Other Factors believed to be associated with differential health outcomes across population groups (i.e., pre-existing conditions, place or state of residence, race/ethnicity/culture/language, immigration, refugee status, occupation, gender identity or sex, religion or belief system, education or literacy level, socioeconomic status, social capital, age, and other factors).[16, 17] Eligible comparators were population groups that did not have the P^2^ROGRESS And Other Factor, or experienced a P^2^ROGRESS And Other Factor to a different degree (e.g., older vs. younger). Factors could be present among a population with or without COVID-19. The infection must have been confirmed by laboratory testing or linked epidemiologically (e.g., household contact). Studies including populations with other pandemic-related infections (e.g., Severe Acute Respiratory Syndrome, Middle East Respiratory Syndrome) were excluded if data specific to COVID-19 cases could not be isolated. We also excluded studies of interventions and where the entire study population had severe disease (e.g., ICU settings).

Any length of follow-up for outcomes of interest was acceptable. Eligible studies reported on at least one primary outcome (i.e., rate of hospitalization, hospital length of stay, severe disease [as defined by study authors; for example, composite outcome of ICU transfer or death], ICU admission and length of stay, need for mechanical ventilation [MV], and mortality [case fatality or all-cause]). In order to prioritize the most rigorous and applicable evidence, we included only prospective and retrospective cohort studies that employed a multivariate analysis and provided results of the independent contribution of P^2^ROGRESS And Other Factors to severe outcomes, while accounting for potential confounders (minimally age and sex). Pre-prints were included only if they were accepted by a peer-reviewed journal; pre-prints that were later published (between the date of the search and manuscript submission) were included. Government reports from hand-searched websites were eligible.

### Study Selection

All records retrieved by the searches were exported to a Microsoft Office Excel (Microsoft Corporation, Redmond, WA) spreadsheet for screening. After piloting the eligibility criteria on a sample of 70 records, one reviewer independently screened records for inclusion by title/abstract, and those deemed to be potentially relevant were assessed by full text. Uncertainties about the inclusion of any full text study were resolved through consultation with a second reviewer.

### Data Extraction

Following a pilot round, one reviewer independently extracted data from each included study into an Excel workbook. We extracted data on (a) population size and demographics, (b) setting, (c) dates of data collection, (d) COVID-19 ascertainment method, (e) co-infections, (f) outcomes reported with definitions for composite outcomes (e.g., severe disease), (f) number of participants analysed, (h) relevant outcome data related to P^2^ROGRESS factors of interest. For both continuous and dichotomous outcomes, we extracted adjusted relative effect sizes (i.e., odds ratio [OR], risk ratio [RR], hazard ratio [HR]) and measures of variability (95% confidence interval [CI]). A second reviewer was consulted in the event of uncertainty about any of the extracted data. Given the expedited approach, we extracted only data that were reported within the included studies and made no attempt to contact authors for missing or unclear data.

### Quality Assessments

To expedite quality assessments, we did not use a formal tool; instead we focused on key variables that were considered to be most relevant to the topic, and that would allow for meaningful stratification of studies by quality. The key variables that we used to assess the quality of the included studies were (a) the extent of adjustment for relevant covariates (i.e., basic adjustment for age and sex, versus more extensive adjustment for numerous potential confounders including comorbidities), (b) follow-up duration and extent of censorship for some outcomes (e.g., ≥2 weeks for mortality), and (c) inappropriate or large exclusions from the study and/or analysis (e.g., missing data on risk factor status or analytical variables). Following assessment of these key variables by a single reviewer, studies without concerns for all three criteria were rated good while others were rated fair. A second reviewer was consulted in the case of uncertainty about the assessment of any individual study.

### Synthesis

Given substantial clinical (e.g., risk factors and/or comparators examined, outcome definitions) and methodological (varying covariates included in the adjusted analyses, different measures of association) heterogeneity, it was not appropriate to pool the studies statistically. Instead, we present a narrative summary of the results across studies for each risk factor. When making conclusions about the association between a P^2^ROGRESS And Other Factor and an outcome, we focused primarily on the magnitude of effect rather than statistical significance, which is heavily dependent on sample size. We categorized associations to be small/unimportant (odds ratio [OR] or risk ratio [RR] ≤1.70), moderate (1.71 to 1.99), large (≥2.00), or very large (≥5.00). [18] When determining the magnitude, we compared findings across all relevant studies and often relied heavily on the findings of the largest and/or good quality studies.

### Certainty of Evidence

The expedited approach to evidence synthesis did not allow for a formal appraisal of the certainty of evidence across studies for each P^2^ROGRESS And Other Factor-Outcome association. Instead, a single reviewer assessed the certainty of the evidence for each association considering relevant components of the Grading of Recommendations Assessment, Development and Evaluation (GRADE) approach: [19, 20] (a) directness in terms of country (presence of universal healthcare) and source population (community sample vs. hospitalized patients), (b) sample size (n<500 considered small) and magnitude of association, (c) study quality, and (d) consistency of associations (in direction and magnitude) across studies. Bodies of evidence started at high certainty[21] and were rated down for weaknesses in any of the aforementioned characteristics. The level of certainty in associations are referred to using the terms ‘uncertain’ (no or very low certainty), ‘may’ (low or some certainty), and ‘probably’ (moderate certainty).[22] At least two other reviewers confirmed the certainty of evidence appraisals, with disagreements resolved by discussion.

## RESULTS

### Characteristics of Studies

Of 3,740 unique records identified by the searches, 949 were screened at full text, and 34 studies that reported on 32 unique populations were included in the review (Figure 1; Supplemental File shows studies excluded by full text, with reasons). [23-56] Three studies conducted in the United Kingdom (UK)[39, 44, 47] used overlapping cohorts from a single medical/research database, and were considered as a single population in the analysis. Another large UK study[56] is likely to also be overlapping with these populations, but the degree of overlap is not known.

**Figure 1.**
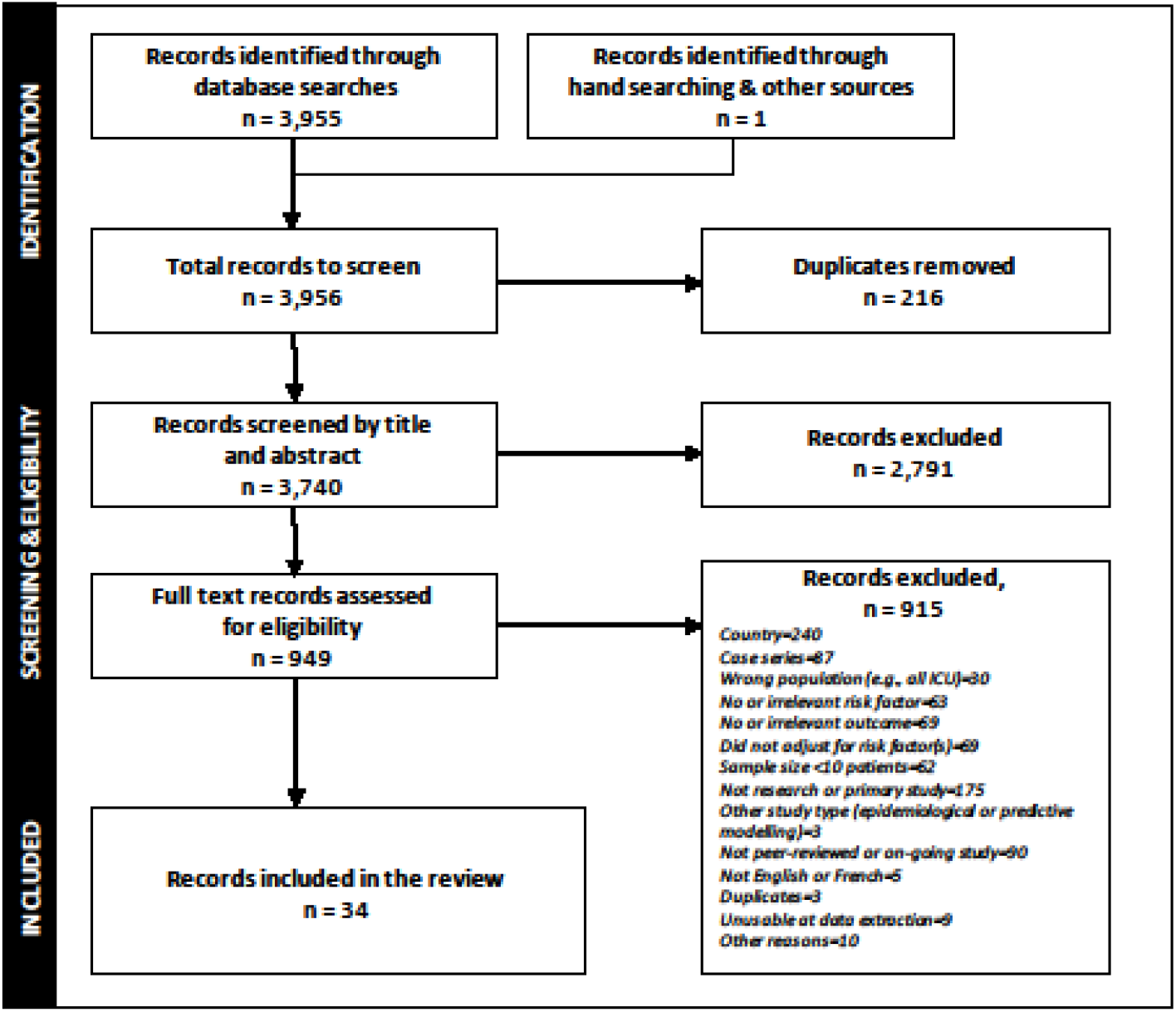
PRISMA flow of study selection.

Table 1 shows the characteristics of the included studies (full details about individual studies in Supplemental File). Briefly, all of the included studies were prospective or retrospective cohorts. The studies were published between 23 April and 6 July 2020, and half (17/34, 50%) reported on populations in the United States.[23, 24, 31, 32, 36-38, 40-43, 45, 46, 49, 51, 53, 54] The remaining countries represented (Italy,[25, 27-30, 35, 50, 55] Spain,[26] UK[33, 39, 44, 47, 48, 52, 56]) all have universal or universal-like healthcare (one study used data from 17 countries). All studies reported on adults, and the overall median was 596 participants (range 44 to 418,794). The mean or median age of the populations studied ranged from 42 to 84 years (32/34 [94%] 54 to 71 years). Most studies (16/34, 47%) examined the association between risk factors and outcomes in a hospitalized population. Studies most commonly reported on the independent association of pre-existing conditions (n=27 studies), gender identity or sex (n=18), and race or ethnicity (n=12) with severe outcomes (most commonly hospitalization, n=9). P^2^ROGRESS And Other Factors not examined in the included studies were immigration or refugee status, religion or belief system, social capital, and substance use disorders. There were also no data specific to pregnant women, indigenous populations, people with disabilities, nor different ages in children.

**Table 1.**
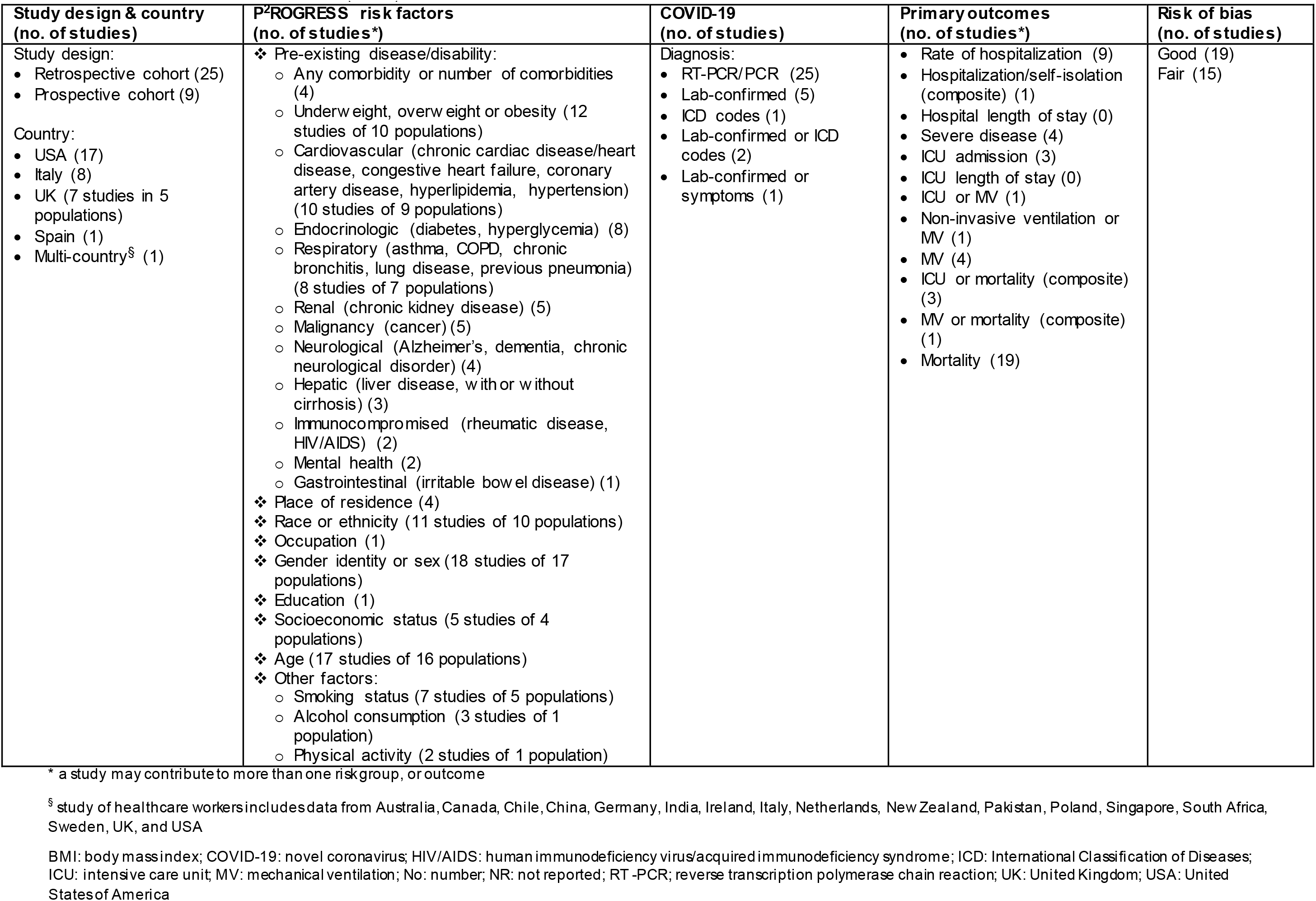
Included studies overview (n=34)

| Study design & country<br>(no. of studies) | P<sup>2</sup>ROGRESS risk factors<br>(no. of studies*) | COVID-19<br>(no. of studies) | Primary outcomes<br>(no. of studies*) | Risk of bias<br>(no. of studies) |
| --- | --- | --- | --- | --- |
| Study design:<br>Retrospective cohort (25)<br>Prospective cohort (9)<br>Country:<br>USA (17)<br>Italy (8)<br>UK (7 studies in 5 populations)<br>Spain (1)<br>Multi-country<sup>§</sup> (1) | ❖ Pre-existing disease/disability:<br>Any comorbidity or number of comorbidities (4)<br>Underweight, overweight or obesity (12 studies of 10 populations)<br>Cardiovascular (chronic cardiac disease/heart disease, congestive heart failure, coronary artery disease, hyperlipidemia, hypertension) (10 studies of 9 populations)<br>Endocrinologic (diabetes, hyperglycemia) (8)<br>Respiratory (asthma, COPD, chronic bronchitis, lung disease, previous pneumonia) (8 studies of 7 populations)<br>Renal (chronic kidney disease) (5)<br>Malignancy (cancer) (5)<br>Neurological (Alzheimer's, dementia, chronic neurological disorder) (4)<br>Hepatic (liver disease, with or without cirrhosis) (3)<br>Immunocompromised (rheumatic disease, HIV/AIDS) (2)<br>Mental health (2)<br>Gastrointestinal (irritable bowel disease) (1)<br>❖ Place of residence (4)<br>❖ Race or ethnicity (11 studies of 10 populations)<br>❖ Occupation (1)<br>❖ Gender identity or sex (18 studies of 17 populations)<br>❖ Education (1)<br>❖ Socioeconomic status (5 studies of 4 populations)<br>❖ Age (17 studies of 16 populations)<br>❖ Other factors:<br>Smoking status (7 studies of 5 populations)<br>Alcohol consumption (3 studies of 1 population)<br>Physical activity (2 studies of 1 population) | Diagnosis:<br>RT-PCR/PCR (25)<br>Lab-confirmed (5)<br>ICD codes (1)<br>Lab-confirmed or ICD codes (2)<br>Lab-confirmed or symptoms (1) | Rate of hospitalization (9)<br>Hospitalization/self-isolation (composite) (1)<br>Hospital length of stay (0)<br>Severe disease (4)<br>ICU admission (3)<br>ICU length of stay (0)<br>ICU or MV (1)<br>Non-invasive ventilation or MV (1)<br>MV (4)<br>ICU or mortality (composite) (3)<br>MV or mortality (composite) (1)<br>Mortality (19) | Good (19)<br>Fair (15) |

\* a study may contribute to more than one risk group, or outcome
<sup>§</sup> study of healthcare workers includes data from Australia, Canada, Chile, China, Germany, India, Ireland, Italy, Netherlands, New Zealand, Pakistan, Poland, Singapore, South Africa, Sweden, UK, and USA
BMI: body mass index; COVID-19: novel coronavirus; HIV/AIDS: human immunodeficiency virus/acquired immunodeficiency syndrome; ICD: International Classification of Diseases; ICU: intensive care unit; MV: mechanical ventilation; No: number; NR: not reported; RT-PCR: reverse transcription polymerase chain reaction; UK: United Kingdom; USA: United States of America

### Study Quality

The majority of studies (19/34, 56%) were rated as good quality[23, 24, 29, 31, 33-36, 40, 42, 45, 46, 48-53, 55] because they adjusted for age, sex, and pre-existing disease in their analysis, had adequate follow-up of outcomes, and few or no missing data. The remaining studies had flaws in one or more of the three domains that we considered to be most important for this review.

### Association Between Risk Factors and Outcomes

Table 2 shows a summary of findings for associations between each reported risk factor and outcomes of interest; all contributing data are shown in the Supplemental File.

**Table 2.**
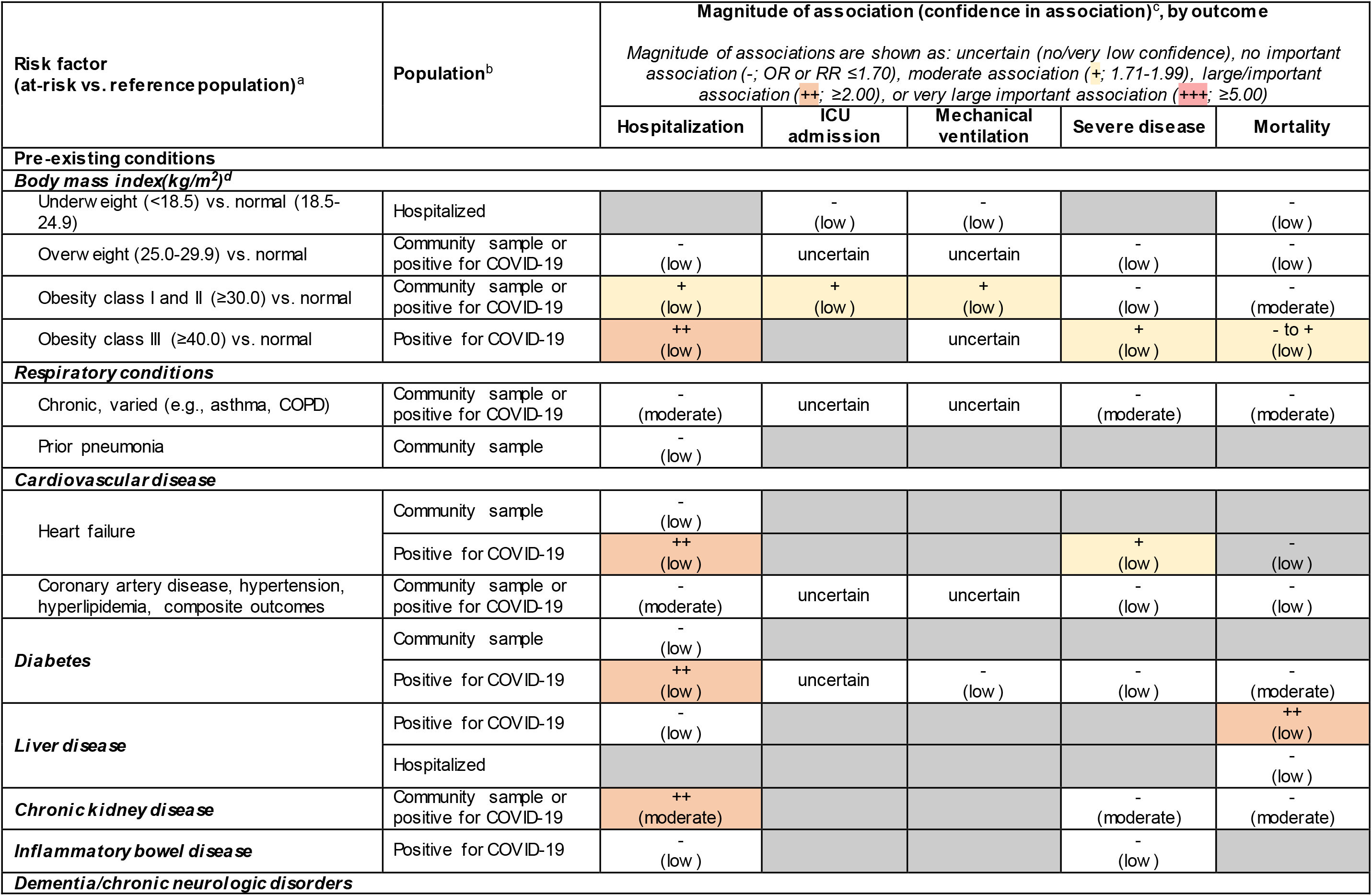
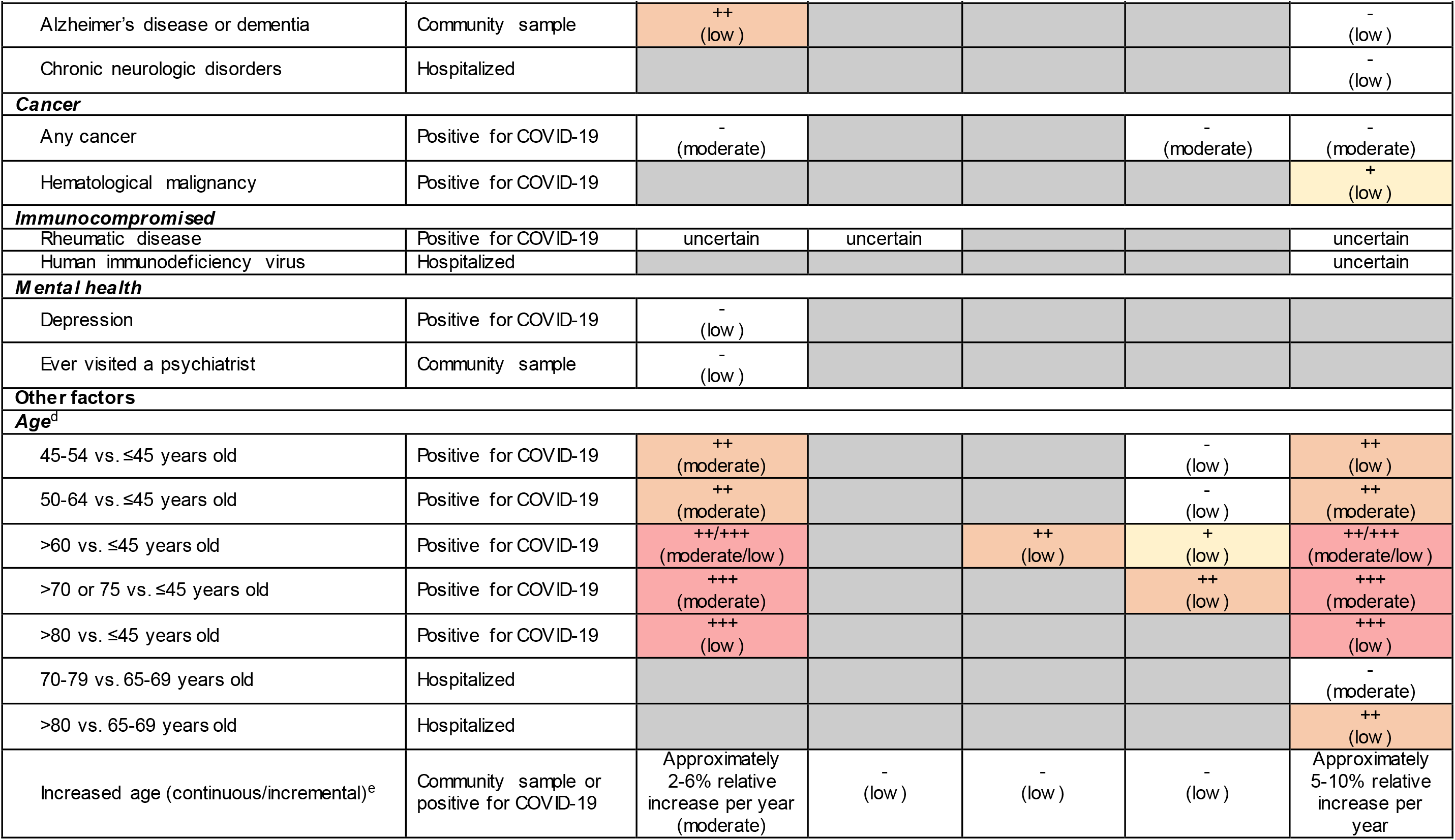
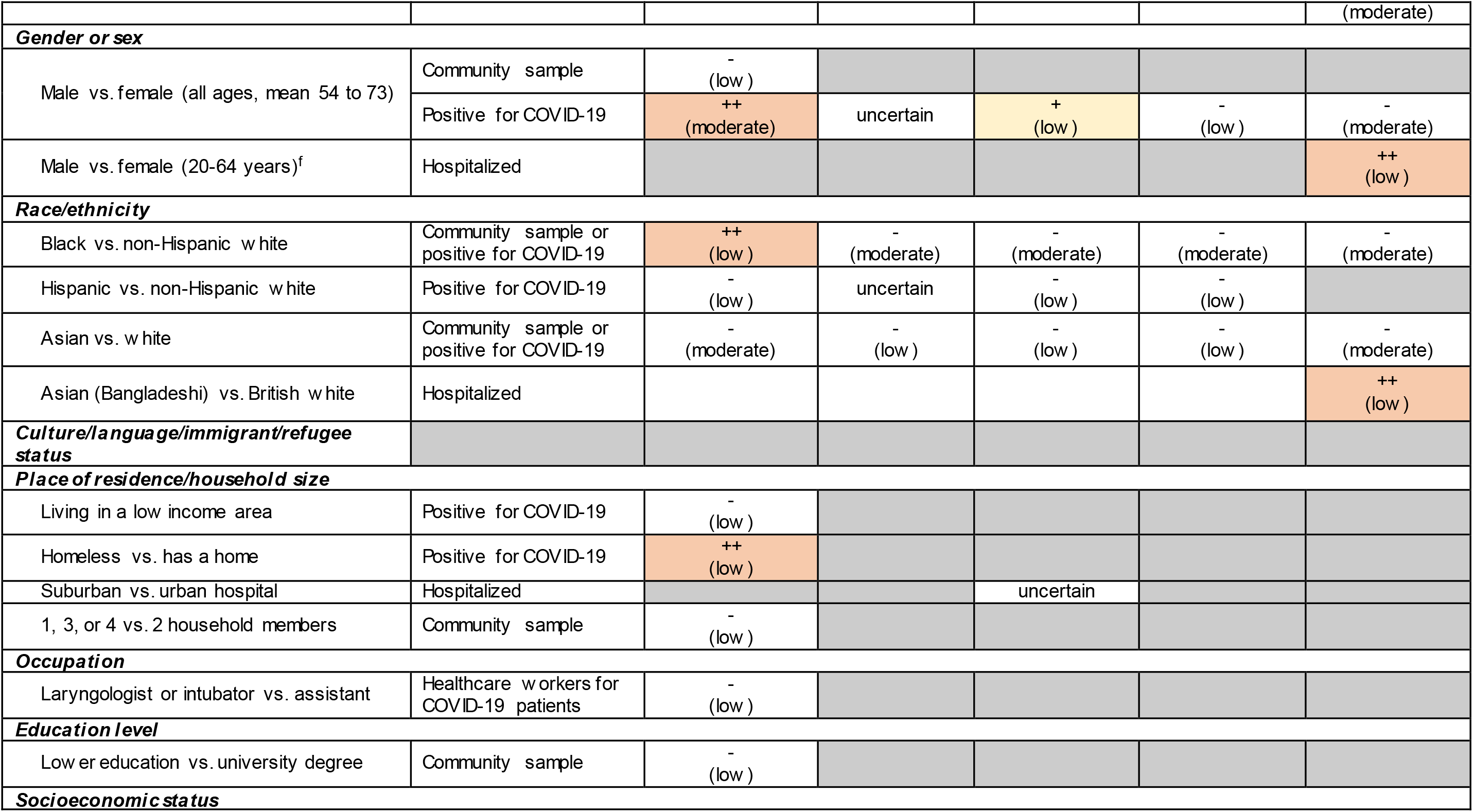
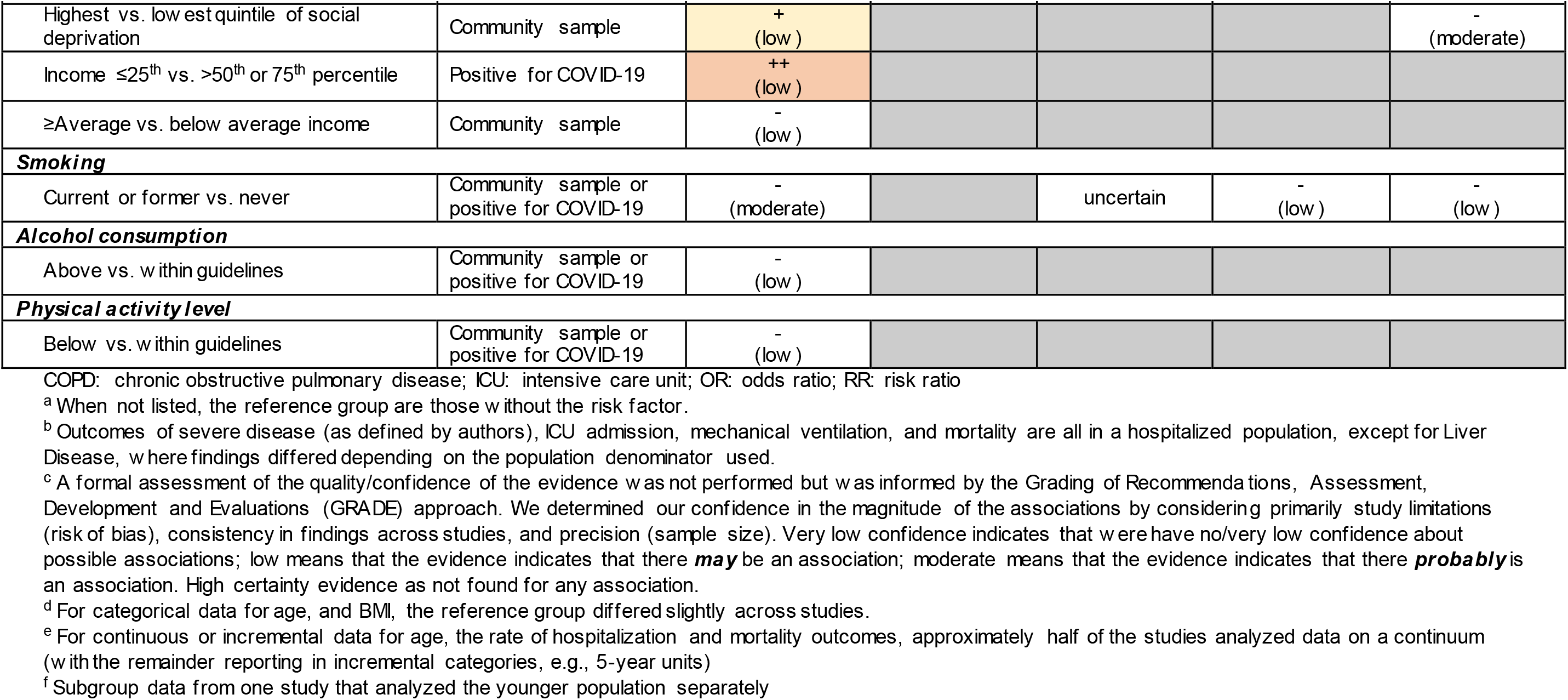
Summary of evidence for associations between risk factors and severe outcomes of COVID-19

| Risk factor<br>(at-risk vs. reference population)<sup>a</sup> | Population<sup>b</sup> | Magnitude of association (confidence in association)<sup>c</sup>, by outcome |  |  |  |  |
| --- | --- | --- | --- | --- | --- | --- |
|  |  | Magnitude of associations are shown as: uncertain (no/very low confidence), no important association (-; OR or RR ≤1.70), moderate association (+; 1.71-1.99), large/important association (++; ≥2.00), or very large important association (+++; ≥5.00) |  |  |  |  |
|  |  | Hospitalization | ICU admission | Mechanical ventilation | Severe disease | Mortality |
| **Pre-existing conditions** |  |  |  |  |  |  |
| **Body mass index(kg/m<sup>2</sup>)<sup>d</sup>** |  |  |  |  |  |  |
| Underweight (<18.5) vs. normal (18.5-24.9) | Hospitalized |  | -<br>(low) | -<br>(low) |  | -<br>(low) |
| Overweight (25.0-29.9) vs. normal | Community sample or positive for COVID-19 | -<br>(low) | uncertain | uncertain | -<br>(low) | -<br>(low) |
| Obesity class I and II (≥30.0) vs. normal | Community sample or positive for COVID-19 | +<br>(low) | +<br>(low) | +<br>(low) | -<br>(low) | -<br>(moderate) |
| Obesity class III (≥40.0) vs. normal | Positive for COVID-19 | ++<br>(low) |  | uncertain | +<br>(low) | - to +<br>(low) |
| **Respiratory conditions** |  |  |  |  |  |  |
| Chronic, varied (e.g., asthma, COPD) | Community sample or positive for COVID-19 | -<br>(moderate) | uncertain | uncertain | -<br>(moderate) | -<br>(moderate) |
| Prior pneumonia | Community sample | -<br>(low) |  |  |  |  |
| **Cardiovascular disease** |  |  |  |  |  |  |
| Heart failure | Community sample | -<br>(low) |  |  |  |  |
|  | Positive for COVID-19 | ++<br>(low) |  |  | +<br>(low) | -<br>(low) |
| Coronary artery disease, hypertension, hyperlipidemia, composite outcomes | Community sample or positive for COVID-19 | -<br>(moderate) | uncertain | uncertain | -<br>(low) | -<br>(low) |
| **Diabetes** | Community sample | -<br>(low) |  |  |  |  |
|  | Positive for COVID-19 | ++<br>(low) | uncertain | -<br>(low) | -<br>(low) | -<br>(moderate) |
| **Liver disease** | Positive for COVID-19 | -<br>(low) |  |  |  | ++<br>(low) |
|  | Hospitalized |  |  |  |  | -<br>(low) |
| **Chronic kidney disease** | Community sample or positive for COVID-19 | ++<br>(moderate) |  |  | -<br>(moderate) | -<br>(moderate) |
| **Inflammatory bowel disease** | Positive for COVID-19 | -<br>(low) |  |  | -<br>(low) |  |
| **Dementia/chronic neurologic disorders** |  |  |  |  |  |  |
|  |  | Magnitude of associations are shown as: uncertain (no/very low confidence), no important association (-; OR or RR ≤1.70), moderate association (+; 1.71-1.99), large/important association (++; ≥2.00), or very large important association (+++; ≥5.00) |  |  |  |  |
|  |  | Hospitalization | ICU admission | Mechanical ventilation | Severe disease | Mortality |
| Alzheimer's disease or dementia | Community sample | ++<br>(low) |  |  |  | -<br>(low) |
| Chronic neurologic disorders | Hospitalized |  |  |  |  | -<br>(low) |
| **Cancer** |  |  |  |  |  |  |
| Any cancer | Positive for COVID-19 | -<br>(moderate) |  |  | -<br>(moderate) | -<br>(moderate) |
| Hematological malignancy | Positive for COVID-19 |  |  |  |  | +<br>(low) |
| **Immunocompromised** |  |  |  |  |  |  |
| Rheumatic disease | Positive for COVID-19 | uncertain | uncertain |  |  | uncertain |
| Human immunodeficiency virus | Hospitalized |  |  |  |  | uncertain |
| **Mental health** |  |  |  |  |  |  |
| Depression | Positive for COVID-19 | -<br>(low) |  |  |  |  |
| Ever visited a psychiatrist | Community sample | -<br>(low) |  |  |  |  |
| **Other factors** |  |  |  |  |  |  |
| **Age<sup>d</sup>** |  |  |  |  |  |  |
| 45-54 vs. ≤45 years old | Positive for COVID-19 | ++<br>(moderate) |  |  | -<br>(low) | ++<br>(low) |
| 50-64 vs. ≤45 years old | Positive for COVID-19 | ++<br>(moderate) |  |  | -<br>(low) | ++<br>(moderate) |
| >60 vs. ≤45 years old | Positive for COVID-19 | ++/+++<br>(moderate/low) |  | ++<br>(low) | +<br>(low) | ++/+++<br>(moderate/low) |
| >70 or 75 vs. ≤45 years old | Positive for COVID-19 | +++<br>(moderate) |  |  | ++<br>(low) | +++<br>(moderate) |
| >80 vs. ≤45 years old | Positive for COVID-19 | +++<br>(low) |  |  |  | +++<br>(low) |
| 70-79 vs. 65-69 years old | Hospitalized |  |  |  |  | -<br>(moderate) |
| >80 vs. 65-69 years old | Hospitalized |  |  |  |  | ++<br>(low) |
| Increased age (continuous/incremental)<sup>e</sup> | Community sample or positive for COVID-19 | Approximately 2-6% relative increase per year<br>(moderate) | -<br>(low) | -<br>(low) | -<br>(low) | Approximately 5-10% relative increase per year |
|  |  | Magnitude of associations are shown as: uncertain (no/very low confidence), no important association (-; OR or RR ≤1.70), moderate association (+; 1.71-1.99), large/important association (++; ≥2.00), or very large important association (+++; ≥5.00) |  |  |  |  |
|  |  | Hospitalization | ICU admission | Mechanical ventilation | Severe disease | Mortality |
|  |  |  |  |  |  | (moderate) |
| **Gender or sex** |  |  |  |  |  |  |
| Male vs. female (all ages, mean 54 to 73) | Community sample | -<br>(low) |  |  |  |  |
|  | Positive for COVID-19 | ++<br>(moderate) | uncertain | +<br>(low) | -<br>(low) | -<br>(moderate) |
| Male vs. female (20-64 years)<sup>f</sup> | Hospitalized |  |  |  |  | ++<br>(low) |
| **Race/ethnicity** |  |  |  |  |  |  |
| Black vs. non-Hispanic white | Community sample or positive for COVID-19 | ++<br>(low) | -<br>(moderate) | -<br>(moderate) | -<br>(moderate) | -<br>(moderate) |
| Hispanic vs. non-Hispanic white | Positive for COVID-19 | -<br>(low) | uncertain | -<br>(low) | -<br>(low) |  |
| Asian vs. white | Community sample or positive for COVID-19 | -<br>(moderate) | -<br>(low) | -<br>(low) | -<br>(low) | -<br>(moderate) |
| Asian (Bangladeshi) vs. British white | Hospitalized |  |  |  |  | ++<br>(low) |
| **Culture/language/immigrant/refugee status** |  |  |  |  |  |  |
| **Place of residence/household size** |  |  |  |  |  |  |
| Living in a low income area | Positive for COVID-19 | -<br>(low) |  |  |  |  |
| Homeless vs. has a home | Positive for COVID-19 | ++<br>(low) |  |  |  |  |
| Suburban vs. urban hospital | Hospitalized |  |  | uncertain |  |  |
| 1, 3, or 4 vs. 2 household members | Community sample | -<br>(low) |  |  |  |  |
| **Occupation** |  |  |  |  |  |  |
| Laryngologist or intubator vs. assistant | Healthcare workers for COVID-19 patients | -<br>(low) |  |  |  |  |
| **Education level** |  |  |  |  |  |  |
| Lower education vs. university degree | Community sample | -<br>(low) |  |  |  |  |
| **Socioeconomic status** |  |  |  |  |  |  |
|  |  | Magnitude of associations are shown as: uncertain (no/very low confidence), no important association (-; OR or RR ≤1.70), moderate association (+; 1.71-1.99), large/important association (++; ≥2.00), or very large important association (+++; ≥5.00) |  |  |  |  |
|  |  | Hospitalization | ICU admission | Mechanical ventilation | Severe disease | Mortality |
| Highest vs. low est quintile of social deprivation | Community sample | +<br>(low) |  |  |  | -<br>(moderate) |
| Income ≤25<sup>th</sup> vs. >50<sup>th</sup> or 75<sup>th</sup> percentile | Positive for COVID-19 | ++<br>(low) |  |  |  |  |
| ≥Average vs. below average income | Community sample | -<br>(low) |  |  |  |  |
| **Smoking** |  |  |  |  |  |  |
| Current or former vs. never | Community sample or positive for COVID-19 | -<br>(moderate) |  | uncertain | -<br>(low) | -<br>(low) |
| **Alcohol consumption** |  |  |  |  |  |  |
| Above vs. within guidelines | Community sample or positive for COVID-19 | -<br>(low) |  |  |  |  |
| **Physical activity level** |  |  |  |  |  |  |
| Below vs. within guidelines | Community sample or positive for COVID-19 | -<br>(low) |  |  |  |  |

COPD: chronic obstructive pulmonary disease; ICU: intensive care unit; OR: odds ratio; RR: risk ratio
<sup>a</sup> When not listed, the reference group are those without the risk factor.
<sup>b</sup> Outcomes of severe disease (as defined by authors), ICU admission, mechanical ventilation, and mortality are all in a hospitalized population, except for Liver Disease, where findings differed depending on the population denominator used.
<sup>c</sup> A formal assessment of the quality/confidence of the evidence was not performed but was informed by the Grading of Recommendations, Assessment, Development and Evaluations (GRADE) approach. We determined our confidence in the magnitude of the associations by considering primarily study limitations (risk of bias), consistency in findings across studies, and precision (sample size). Very low confidence indicates that we have no/very low confidence about possible associations; low means that the evidence indicates that there **may** be an association; moderate means that the evidence indicates that there **probably** is an association. High certainty evidence as not found for any association.
<sup>d</sup> For categorical data for age, and BMI, the reference group differed slightly across studies.
<sup>e</sup> For continuous or incremental data for age, the rate of hospitalization and mortality outcomes, approximately half of the studies analyzed data on a continuum (with the remainder reporting in incremental categories, e.g., 5-year units)
<sup>f</sup> Subgroup data from one study that analyzed the younger population separately

There was low or moderate certainty of evidence for important/large associations with increased risk of hospitalization in people having confirmed COVID-19, for the following risk factors: obesity class III (body mass index ≥40 kg/m^2^; 1 study, n=5,297),[49] heart failure (2 studies, n=6,331),[23, 49] diabetes (2 studies, n=6,331),[23, 49] chronic kidney disease (confirmed COVID-19 or community sample; 2 studies, n=424,073),[47, 49] dementia (1 study, n=418,794),[47] age over 45 years (vs. 45 or younger; 2 studies, n=6,331),[23, 49] male gender (3 studies, n=3,812),[23, 49, 51] black race/ethnicity (vs. non-Hispanic white; confirmed COVID-19 and community samples, 5 studies in 4 populations, n=428,606), [23, 44, 47, 49, 51] homelessness (1 study, n=1,052),[23] and low income (<25^th^ vs. >50^th^ percentile; 1 study, n=1,052).[23] Age over 60 and over 70 years may be associated with important increases in the rate of mechanical ventilation (1 study, n=486)[40] and severe disease (1 study, n=2,725), [49] respectively.

For mortality, important associations with increased risk may exist for liver disease (2 studies, n=20,597),[33, 53] Bangladeshi ethnicity (vs. British white; 1 study, n=130,091), [56] and age over 45 years (vs. <45 years; 3 studies, n=87,819).[33, 49, 56] The data were somewhat inconsistent for gender, with most studies showing moderate certainty of no important effect, but one large fair quality study (n=130,091)[56] from the UK that stratified its analysis by age showed that hospitalized males aged 20-64 years (but not older) may be at about two-fold increased risk of mortality compared to females. Associations with hospitalization and mortality may be very large for those aged over 60 years (2 studies, n=6,331 for hospitalization;[23, 49] 3 studies, n=24,163 for mortality[33, 41, 49]) and are probably very large for those over 70 years (2 studies, n=6,331 for hospitalization; [23, 49] 2 studies, n=22,858 for mortality[33, 49]). One study (n=63,094)[56] directly compared subgroups of older adults, showing that compared to those aged 65-69 years, there may be no important increased risk of mortality among hospitalized adults aged 70-79 years, but risk may increase about 2-fold for those 80 years and older. Studies treating age on a continuum or across small increments consistently found that risks for hospitalization and mortality increased with increasing age (e.g., approximately 2-6% and 5-10% relative increase in risk per year) (3 studies in 2 populations, n=422,275 for hospitalization; [44, 47, 51] 11 studies, n=6,877 for mortality).[25-27, 31, 35, 38, 45, 46, 48, 51, 55]

Moderate associations may exist for increased risk of mechanical ventilation (4 studies, n=1,559)[38, 40, 42, 46] and ICU admission (2 studies, n=873),[38, 42] and severe disease (1 study, n=2,725)[49] with obesity (body mass index ≥30 or 40 kg/m^2^); severe disease with heart failure (1 study, n=2,725);[49] mortality with haematological malignancy (1 study, n=1,183);[52] mechanical ventilation with male gender (4 studies, n=881);[27, 40, 42, 46] and hospitalization with social deprivation (highest vs. lowest quintile; 1 study, n=340,996).[44]

There was moderate certainty evidence for no important increase in risk of hospitalization with chronic respiratory conditions (4 studies in 3 populations, n=425,125), [23, 44, 47, 49] cardiovascular disease apart from heart failure (i.e., coronary artery disease, hypertension, hyperlipidaemia; 4 studies in 3 populations, n=425,125),[23, 44, 47, 49] non-specific cancer (2 studies, n=6,331), [23, 49] Asian race/ethnicity other than Bangladeshi (vs. non-Hispanic white; 3 studies in 2 populations, n=424,073), [44, 47, 49] and current or former smoking (5 studies in 3 populations, n=425,125). [23, 39, 44, 47, 49] Additionally, there was moderate certainty evidence for no important increase in severe disease with chronic respiratory conditions (1 study, n=2,725), [49] chronic kidney disease (2 studies, n=2,922),[24, 49] nonspecific cancer (2 studies, n=2,769), [29, 49] and Black race/ethnicity (vs. nonHispanic white; 2 studies, n=3,030);[36, 49] and no important increase in risk of mortality with obesity (body mass index ≥30 kg/m^2^; 6 studies, n=8,716),[35, 38, 43, 46, 49, 51] chronic respiratory conditions (4 studies, n=23,315),[31, 33, 46, 49] diabetes (4 studies, n=23,315), [31, 33, 46, 49] chronic kidney disease (3 studies, n=23,058), nonspecific cancer (3 studies, n=24,041),[33, 49, 52] male gender (9 studies, n=27,875),[25-27, 31, 33, 35, 46, 49, 51] Black (5 studies, n=135,418)[38, 48, 49, 51, 56] or Asian race/ethnicity (vs. non-Hispanic white; 3 studies, n=4,015),[38, 48, 49] and social deprivation (lowest vs. highest quintile; 1 study, n=130,091).[56] Overall, there were few data for the ICU and mechanical ventilation outcomes.

## DISCUSSION

Responding to an urgent need for empiric evidence to inform decision-making on Canada’s immunization strategies,[7] in this rapid review we synthesized studies employing a multivariate analysis to ascertain potential independent associations between “P^2^ROGRESS And Other Factors” and severe outcomes of COVID-19. Among 22 potential risk factors examined across the included studies, the most important risk factors (i.e., those associated with large/important increased risk; OR or RR ≥2.0) for hospitalization among those with confirmed COVID-19 were several pre-existing chronic health conditions (obesity class III, heart failure, diabetes, chronic kidney disease [community sample or with COVID-19], dementia [community sample]), older age (>45 years vs. younger), male gender, Black race/ethnicity (community sample or with COVID-19), homelessness, and low income (≤25^th^ vs. >50^th^ percentile). Liver disease may be associated with a large increased risk of mortality among people with COVID-19 and advancing age (>45 years vs. younger) and Bangladeshi ethnicity (vs. British white) are likely to be associated with a large increased risk of mortality among hospitalized patients. There is evidence to suggest that male gender may increase risk of mortality among younger (20-64 years), but not older men.

Among the factors identified as increasing risk of severe outcomes, age seemed to be the most influential; adults older than 60 years may have at least 5 times increased odds of hospitalization and mortality from COVID-19 compared to those aged less than 45 years. This increased risk appears to magnify at least to some degree even for those older than 60 years, with those aged over 80 years having double the mortality risk of those aged 65-69 years. Though we focused the review on better quality studies that minimally controlled for age and sex, the strength of certain associations should be interpreted cautiously because there are likely to be multiple unmeasured confounders that have not been accounted for. For example, studies reporting on associations between outcomes and age did not adjust for nursing home residency, and studies examining race did not account for occupation, which may be an important confounder influencing susceptibility to the infection. [56] In addition, it is important to be aware that criteria for COVID-19 testing and hospitalization may differ by place and time, but it is difficult to predict how this may have impacted the findings. In general, many studies conducted testing based on symptoms and the evidence is likely most applicable to these populations. The evidence for mechanical ventilation, ICU admission, and severe disease outcomes was relatively sparse, and we located no evidence meeting our publication date and inclusion criteria to inform the impact of immigration or refugee status, religion or belief system, social capital, substance abuse disorders, pregnancy, indigenous identity, living with a disability, nor differing levels of risk among children in various age groups.

The findings of this rapid review will be used to populate the Equity Matrix of NACI’s Ethics, Equity, Feasibility, and Acceptability Framework,[9] which will be a part of a suite of considerations for informing the development of NACI recommendations on priority pandemic immunization strategies when initial COVID-19 vaccine supply is limited. NACI will be using the results of this rapid review and their current understanding of the epidemiology of COVID-19 in Canada to identify distinct inequities associated with COVID-19, potential reasons for these inequities, and suggested interventions to reduce inequities and improve access to vaccine when it becomes available. The Equity Matrix applied to COVID-19 with evidence to-date can be found elsewhere.[11]

### Strengths and Limitations

The expedited methods used in this review allowed for a rapid but comprehensive synthesis of the highest quality evidence available on multiple risk factors associated with severe COVID-19 outcomes that is applicable to OECD countries. Generalizations to other countries should be made with caution, as high risk groups in these populations may differ. We excluded studies only examining patients with severe COVID-19 (i.e., in ICU settings), and therefore our findings for mechanical ventilation and mortality are applicable to people with COVID-19 or in general populations, but not necessarily all those with severe infection. Most studies of patients in the ICU setting that we located were relatively small and descriptive in nature, such that many would have been excluded due to lack of adjustment or only have been able to provide low or very low certainty evidence due to their lack of precision. As described previously, many available studies do not control for any important confounding variables which limited the number of studies and risk factors included in this review. Given the rapid emergence of new evidence on the topic, potential associations (or lack of association) for which only low or very low certainty of evidence is available should continue to be reviewed as new primary research is published. There is a need for high quality primary research (accounting for multiple confounders) to better understand the level of risk that might be associated with immigration or refugee status, religion or belief system, social capital, substance abuse disorders, pregnancy, indigenous identity, living with a disability, and differing levels of risk among children in various age groups.

## Data Availability

Data sharing statement: All data used in this review are available within the manuscript and accompanying supplemental files.

## ACKNOWLEDGMENTS

We would like to thank the National Advisory Committee on Immunization (NACI) High Consequence Infectious Disease Vaccine Working Group (Caroline Quach, Shelley Deeks, Yen Bui, Kathleen Dooling, Robyn Harrison, Kyla Hildebrand, Michelle Murti, Jesse Papenburg, Robert Pless, Nathan Stall, and Stephen Vaughan) for their contributions to the project. We also thank Liz Dennett (MLIS) for conducting the Medline search, and Karyn Crawford for assisting with article retrieval.

**ABBREVIATIONS**
COVID-19: Novel coronavirus disease 2019
ICU: Intensive care unit
NACI: National Advisory Committee on Immunization
OECD: Organisation for Economic Co-operation and Development
OR: Odds ratio
P2ROGRESS: Pre-existing disease or disability, place of residence, race, ethnicity, culture, language, immigrant/refugee status, occupation, gender, religion/belief system, education, socioeconomic status, social capital, age, and other factors
RR: Risk ratio

## Contributor and guarantor information

AW, JP, AB, BV and LH contributed to the conception and design of the study. AW, JP, SG, SR, AB, and BV contributed to the screening of eligible studies. AW, SG, SR, and AB contributed to the acquisition of data. AW, JP, MG, BV and LH contributed to the synthesis and interpretation of data. AW, JP, MG, drafted the manuscript. AW, JP, MG, SG, SR, AB, BV, and LH revised the manuscript for important intellectual content. All authors approved the manuscript for submission.

## Data sharing statement

All data used in this review are available within the manuscript and accompanying supplemental files.

